# Muscle fatigue during spinal stimulation and resistive ankle exoskeleton use in children with cerebral palsy

**DOI:** 10.1101/2025.08.23.25334289

**Authors:** Charlotte R. DeVol, Victoria M. Landrum, Siddhi R. Shrivastav, Heather A. Feldner, Kristie F. Bjornson, Chet T. Moritz, Katherine M. Steele

## Abstract

Evaluating fatigue during rehabilitation can help prevent overexertion to improve motor learning. The purpose of this study was to quantify how walking with transcutaneous spinal cord stimulation (tSCS) impacts muscle fatigue during treadmill training with and without a resistive ankle exoskeleton (Exo) in children with cerebral palsy (CP). Nine children with CP (4-14 years old) participated in four walking conditions: (1) No Device, (2) tSCS only, (3) Exo only, and (4) tSCS+Exo. Plantarflexor maximum voluntary contraction (MVC) was performed before and after each condition. Soleus amplitude root mean square (RMS) and median frequency (MDF) from electromyography recordings were used as biomarkers for fatigue. Participants only showed signs of soleus fatigue in the first five minutes of the Exo only condition with increased RMS (p < 0.001) and decreased MDF (p < 0.001). After ten minutes of walking, there was a slight decrease in MDF in the No Device and tSCS only conditions (p = 0.002), but no change in RMS. There were no significant differences in changes in MVC force, RMS, or MDF between conditions. Walking with tSCS may reduce the impact of the Exo on soleus fatigue for children with CP.

## Introduction

One of the main challenges among ambulatory people with cerebral palsy (CP) is fatigue in daily life. Individuals with CP use two to three times the energy of nondisabled peers to walk^1–3^. Fatigue limits mobility for both children and adults with CP, affecting their ability to perform tasks of daily living and interact with peers^4–6^. One component of fatigue is muscle fatigue, defined as a reduction in force output^7,8^. Prior to an observable decrease in force output, there are myoelectric manifestations of muscle fatigue. This includes a decrease in the conduction velocity of muscle fibers, which can be quantified as a simultaneous decrease in the frequency and increase in amplitude of electromyographic (EMG) recordings^9^. When performing repetitive tasks, such as walking, the muscles of children with CP fatigue faster than their peers^10–12^. Changes in muscle properties and reductions in the ability to activate all available muscle fibers may make children with CP more likely to rely on weaker, high endurance muscle fibers compared to peers and, thus, make them more prone to muscle fatigue^13^.

Considering muscle fatigue is important when developing rehabilitation interventions. When physical training, such as walking, is performed with rest breaks to prevent over-exertion, the nervous system is more adaptable to learning new information, a process known as neuroplasticity^14^. Neuroplasticity is key for children with CP who are learning new tasks during physical therapy. High muscular effort can cause sustained muscle fatigue during a training session and may reduce neuroplasticity and performance^14,15^. Despite the importance of muscle fatigue on neuroplasticity, fatigue is often not quantified during the development or evaluation of rehabilitation interventions. Understanding fatigue during training can provide useful information for determining optimal session length and level of exertion during an intervention, thus helping develop more effective and individualized training.

Several new rehabilitation interventions have recently been introduced for children with CP that may influence muscle fatigue, neuroplasticity, and function. Exoskeletons that provide ankle resistance during push-off have improved walking speed and planarflexor control^16–18^. However, resistance applied during gait training may lead to more muscle fatigue compared to standard gait training. Understanding the effects of a resistive ankle exoskeleton on muscle fatigue may inform treatment session length or resistance levels to optimize conditions for neuroplasticity. Similarly, neuromodulation may also be a viable tool for reducing muscle fatigue and optimizing performance. Specifically, transcutaneous spinal cord stimulation (tSCS) combined with physical training, such as gait training, has recently shown promising improvements in muscle co-contraction, gross motor function, spasticity, and gait kinematics for children with CP^19–24^. Applying tSCS during walking can reduce muscle co-contraction and improve coordination between hip and knee motion during gait^21^. Through improved coordination, tSCS may reduce muscle demand during training to mitigate fatigue and improve conditions for neuroplasticity. Understanding the impact of tSCS on muscle fatigue may assist in its translation into clinical care.

The purpose of this study was to quantify muscle fatigue before, during, and after gait training with and without tSCS and a resistive ankle exoskeleton (Exo) targeting plantarflexor muscles for children with CP. We evaluated muscle activity before, during, and after walking on a treadmill under four conditions: No Device, tSCS only, Exo only, and tSCS + Exo. Compared to walking with No Device, we hypothesized that the Exo would increase muscle fatigue. We further hypothesized that the combination of tSCS with the Exo would reduce the rate of fatigue compared to the Exo alone.

## Methods

### Participants

Nine children with CP were recruited to evaluate gait training with tSCS and an Exo (Table 1). Prior to enrollment, informed age-appropriate assent and consent were obtained from participants and their caregivers, respectively. All visits were completed at the University of Washington. This study was approved by the University of Washington Human Subjects Division (IRB identifier: STUDY00014877) and registered at ClinicalTrials.gov (NCT05520359; registered on 25 August 2022). The study methods were performed in accordance with the approved protocol and University of Washington Institutional Review Board guidelines and regulations. Individuals were eligible to participate if they were ambulatory, Gross Motor Function Classification System levels I-II, had spastic CP, had no prior selective dorsal rhizotomy, had not undergone a lower extremity surgery or botulinum toxin injections in the past year, did not have uncontrolled seizures, were able to follow 2-3 step commands, and had no implants that might interfere with electrical stimulation. Informed consent was obtained from all participants’ legal guardians for the use of any photos within the manuscript that show a participant.

**Table 1.** Participant Characteristics:

| ID | Sex | Age Range (years) | Height (cm) | Weight (kg) | Spastic CP Diagnosis | Side | GMFCS Level | Speed (m/s) | tSCS Amplitude (mA) |  |
| --- | --- | --- | --- | --- | --- | --- | --- | --- | --- | --- |
|  |  |  |  |  |  |  |  |  | T11 | L1 |
| P01 | M | 10-14 | 169 | 64.3 | Unilateral | R | I | 1.2 | 30 | 20 |
| P02 | M | 0-4 | 109 | 17.2 | Bilateral | L | I | 0.8 | 35 | 25 |
| P03 | M | 10-14 | 155 | 38.6 | Bilateral | R | II | 1.0 | 20 | 30 |
| P04 | M | 15-19 | 160 | 48.2 | Bilateral | L | II | 0.9 | 35 | 25 |
| P05 | F | 5-9 | 117 | 20.5 | Bilateral | R | II | 0.6 | 10 | 20 |
| P06 | F | 5-9 | 113 | 19.5 | Bilateral | R | II | 0.7 | 10 | 15-20 |
| P07 | F | 10-14 | 157 | 51.3 | Bilateral | L | I | 1.0 | 20 | 30 |
| P08 | M | 15-19 | 163 | 58.3 | Bilateral | L | II | 1.1 | 30 | 20 |
| P09 | M | 10-14 | 135 | 39.0 | Unilateral | R | II | 0.8 | 10 | 20 |

ID = participant identifier; Side = more affected side based on the side with more spasticity at baseline and parent reports; GMFCS = Gross Motor Function Classification System; The tSCS amplitude applied to T11 = thoracic spinous process 11, L1 = lumbar spinous process 1.

### Experimental Protocol

Participants completed four visits where they walked with or without an Exo and tSCS (Figure 1). These walking conditions included: No Device, tSCS only (tSCS), resistive exoskeleton (Exo), or the combination of tSCS and Exo (tSCS+Exo). P06 and P07 did not complete the tSCS+Exo visit due to a change in eligibility. Participants walked on the treadmill at a preferred speed that was determined at the first visit and held constant at subsequent visits. Walking conditions were performed on an inground treadmill (Bertec, Columbus, OH) on separate days at least five days apart in a pseudo-randomized order. Rest breaks were provided as needed with a maximum of two rest breaks during each session.

**figure 1.**
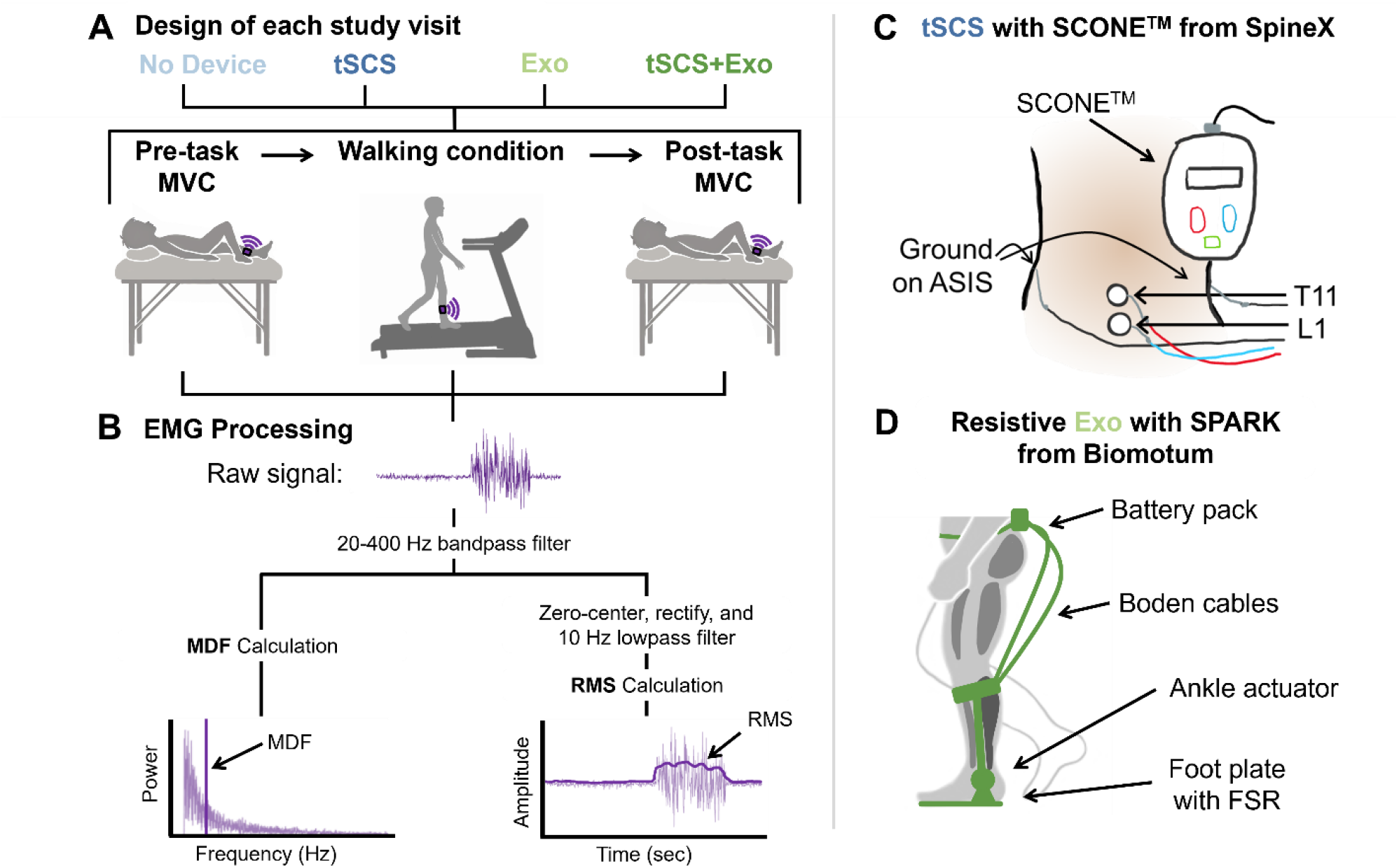
overview of study design, data processing, and devices evaluated. a) participants completed four study visits walking for a maximum of 20 minutes with: no device, spinal stimulation (tscs), a resistive ankle exoskeleton (exo), and both tscs and exo (tscs+exo). participants performed a maximum voluntary contraction (mvc) of the plantarflexor muscles while lying in a supine position before and after walking. soleus muscle activity was recorded during each mvc and walking condition. b) processing pipeline for electromyography (emg) data to calculate median frequency (mdf) and amplitude root mean square (rms) as biomarkers of muscle fatigue. c) placement of electrodes for the spinal cord neuromodulator (sconetm) from spinex. d) the spark exoskeleton by biomotum provided a torque at the ankle to resist during push-off. fsr = force sensitive resistor.

For the tSCS and tSCS+Exo visits, tSCS was administered via the Spinal Cord Neuromodulator (SCONE™, SpineX, Inc.) following previously reported protocols^21^. Stimulation was applied using adhesive gel electrodes placed just below the T11 and L1 spinous processes using 3.2 cm round electrodes. The ground electrodes were 5.1 x 8.6 cm rectangular electrodes placed over the anterior superior iliac spine (Figure 1C). The amplitude for the subthreshold stimulation was determined foreach individual at their first visit using tSCS and held constant at subsequent visits, with the exception of P06 who needed a slight adjustment of tSCS due to comfort during their first use of tSCS (Table 1). The amplitude of tSCS was determined based on children’s self-reported ease of walking, sensation beneath the cathodes, and a clinical interventionist’s observation of gait quality and participant’s behavior within the first ten minutes of use. The tSCS was then turned off and a break was provided until walking began. The amplitude of tSCS was also lowered or turned off during seated breaks and turned back on during walking, as sitting can change the sensation at the site of stimulation compared to standing^25^.

For the Exo and tSCS+Exo walking conditions, we used the SPARK exoskeleton by Biomotum (Portland, OR) to provide resistance at the ankle during the push-off phase of gait^17^. The SPARK is an untethered exoskeleton with a waist strap that carries the battery pack, Bowden cables that connect the battery and motor to an ankle actuator, and foot plates inside the users’ shoe. The SPARK records timing of gait events from a force sensitive resistor in the foot plate and uses this information to time when it applies a torque at the ankle to resist push-off (Figure 1D). The ankle resistance torque of the Exo was set at 12% of each participant’s bodyweight, similar to previous studies^16,17,26^. The SPARK pairs with a mobile application to control the level of resistance, which can also provide audiovisualfeedback on the torque applied at the ankle. The audiovisual feedback feature was not used during the first five minutes of walking with the Exo. Data after this period, when the audiovisual feedback was used, was excluded from this analysis because it may impact soleus muscle activity^26^.

During all walking conditions, electromyography (EMG) data of the soleus muscle were recorded at 2000 Hz (Delsys, Inc., Natick MA). We focused on the soleus muscle because the Exo is designed to target the plantarflexor muscles, and other plantarflexors are not accessible to record with surface EMG with the Exo’s calf cuff in place^17^. We analyzed the more-affected lower limb for all participants, defined as the side of the body with more spasticity. Before (pre) and after (post) each walking condition, maximum voluntary contraction (MVC) of the more-affected plantarflexors were obtained using an isometric manual muscle test with a hand-held dynamometer (Activbody Inc., San Diego, CA). Participants laid supine and were instructed to keep their knees fully extended while plantarflexing their foot as strongly as possible for 5 seconds against a researcher providing resistance with the hand-held dynamometer^18^. Participants were given visual feedback of their force value output and instructed to raise the number as high as possible and continue pushing until time was up. The MVC was repeated three times before and after each walking condition and EMG data of the soleus muscle were recorded throughout and averaged across the three trials. Trials were dropped if there was an outlier in force value, as judged visually by the research team during a session, and an extra MVC was taken to include three total MVC’s before and after each walking condition. P06 did not complete the MVC for any walking conditions.

### Data Analysis

Soleus muscle median frequency (MDF) and root mean square (RMS) were used as biomarkers for myoelectric manifestations of muscle fatigue^9,27^. EMG data for walking conditions were analyzed using a custom MATLAB script (MathWorks, Natick, MA). Soleus EMG signals were bandpass filtered from 20 – 400 Hz using a 4th order Butterworth filter. Soleus firing rate was quantified by segmenting bandpass filtered soleus EMG using a Gabor transform moving window centered at 15-second intervals and calculating the MDF within each window^10,28^. The Gabor transform filters EMG data in the time domain using a Gaussian function,

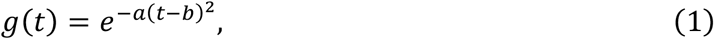

where *a* = −0.01 is the window width of the filter, tis the time, and *b* = 15 seconds, the distance between centers of the moving window. The MDF was calculated using the built-in MATLAB function, medfreq(). EMG data were then zero-centered, rectified, and low-pass filtered (4th order Butterworth; 10 Hz; Figure 2) before calculating RMS amplitude in 15-second intervals^27^. Soleus firing rate and RMS were normalized to the calculated firing rate and peak activation of the first 15 seconds of walking, respectively, so reported values are a percent of the start value.

**Figure 2.**
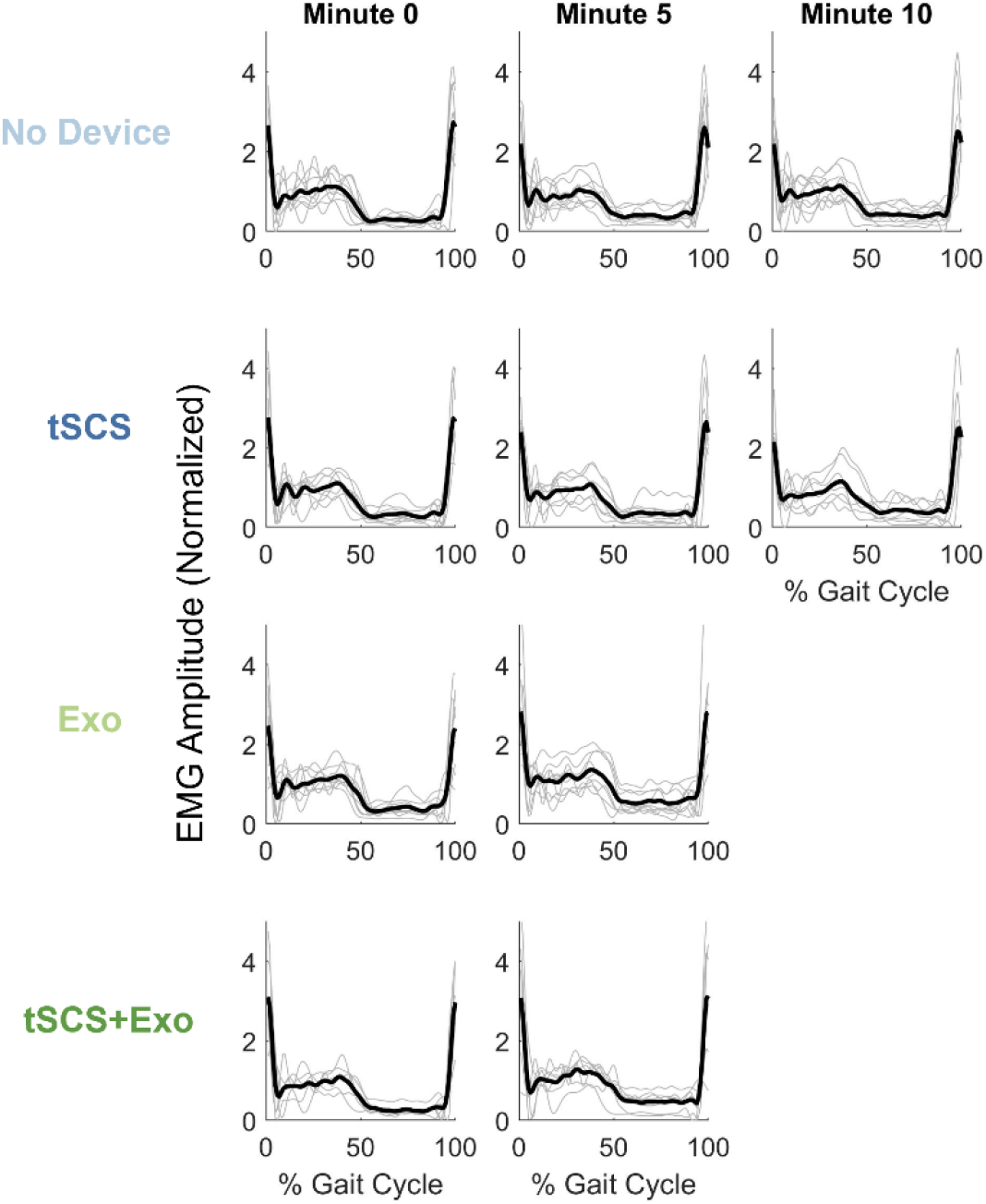
Soleus EMG over the gait cycle for each walking condition: No Device, tSCS only, Exo Only, and tSCS+Exo. Each plot shows the gait cycles averaged over a 15-second duration at minute zero, five, and ten. Thin grey lines are for each participant and the thick black lines are the average across participants. The vertical axis is the EMG amplitude normalized to the RMS over the first 15-seconds.

During the five second MVC task, EMG data were also recorded. The same EMG analyses described above were repeated, except that MDF and RMS calculations were made over the entire five second window.

### Statistical Analysis

We used linear mixed effects models (LMM) to quantify the rate of change of EMG RMS and MDF across five minutes of treadmill walking for all four walking conditions: No Device, tSCS only, Exo only, and tSCS+Exo. Only the first five minutes were used for all four conditions because after this point, participants began taking breaks or using audiovisual feedback in the conditions that included Exo, potentially altering muscle fatigue in a nonlinear manner^26^. Use of an LMM was chosen since all walking conditions contained the same participants. The equations for the EMG RMS and MDF LMMs were:

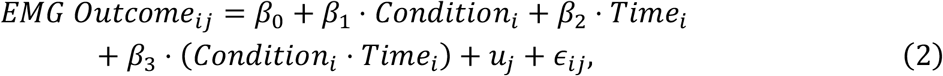

where *EMG Outcome ij* is the observed value of EMG RMS or MDF for the *i*-th observation of the𝑗-th subject. These equations had the best model fit under the assumption that both walking condition and time influence fatigue rate. We also extended the LMM to included 10 minutes of treadmill walking for tSCS and No Device conditions, during which no participants took rest breaks. Time was measured in seconds, resulting in the fixed effects units as percent per second (%/s) and reported with the 95-percent confidence intervals in brackets.

We compared percent change in force output, EMG RMS, and EMG MDF during the post-post MVC task recorded before and after each walking condition. Data sets were not normally distributed, as tested by the Kolmogorov-Smirnov test and a Kruskal-Wallis test was used to measure significant differences between walking conditions. All statistical tests were performed in MATLAB.

## Results

### Fatigue During Five Minutes of Treadmill Walking: All Conditions

Muscle fatigue, quantified as an increase in RMS and a decrease in MDF, occurred only during the first five minutes of the Exo condition (Figure 3). Walking with No Device for five minutes did not cause a significant change in either RMS or MDF (p = 0.67 and 1.0, respectively). Walking with the Exo, there was a significant increase in RMS (*𝛽*_3_ = 0.066%/s [0.029, 0.10]) and decrease in MDF (*𝛽*_3_ = -0.081 %/s [-0.11, -0.049])compared to walking with No Device (p<0.001). Walking with tSCS only or Exo + tSCS had no effect on RMS (tSCS p = 0.40; Exo + tSCS p = 0.43) or MDF (tSCS p = 0.46; Exo + tSCS p = 0.89) compared to walking with No Device.

**Figure 3.**
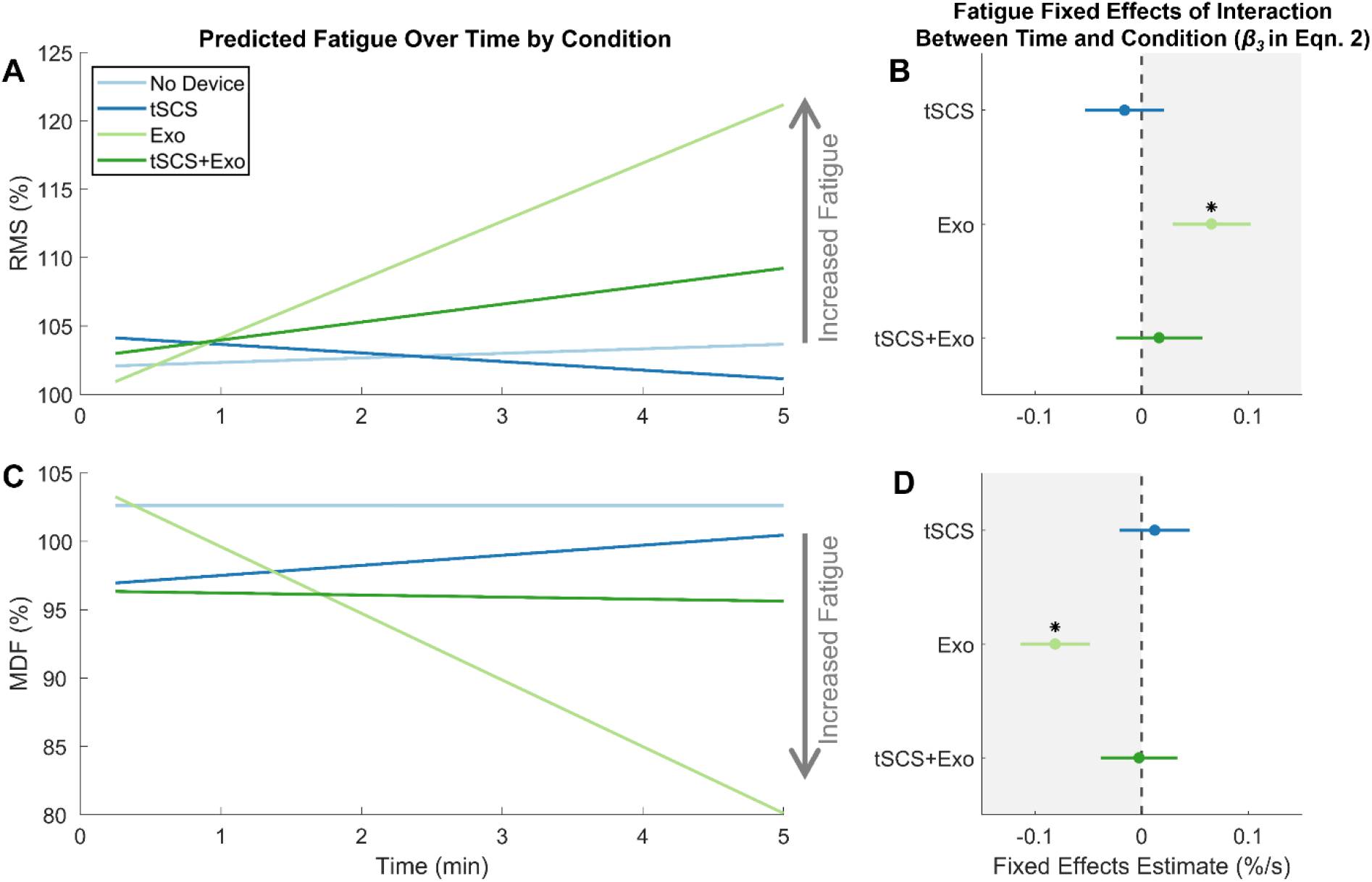
Myoelectric manifestations of muscle fatigue during five minutes treadmill walking under the four different conditions: No Device, tSCS only, Exo only, and tSCS+Exo. A) The linear mixed effects model (LMM) for RMS of the EMG signal normalized to the peak RMS during the first 15 seconds of each condition. B) The RMS fixed effects (𝛽_3_ in Eqn. 2) of the interaction between time and condition with respect to the No Device condition show a significant increase with Exo only. C) The LMM for MDF of the EMG signal, normalized to the MDF in the first 15 seconds of each condition. D) The MDF fixed effects (𝛽_3_ in Eqn. 2) of the interaction between time and condition with respect to the No Device condition show a significant decrease with Exo only. Areas indicating an increase in fatigue relative to No Device are indicated with grey arrows and shaded boxes. * significant difference relative to No Device condition (p < 0.05).

### Fatigue During Ten Minutes of Treadmill Walking: No Device and tSCS Only

We also quantified changes in RMS and MDF during ten minutes of consecutive walking with No Device and tSCS only to see if longer trials would induce muscle fatigue (Figure 4).

**Figure 4.**
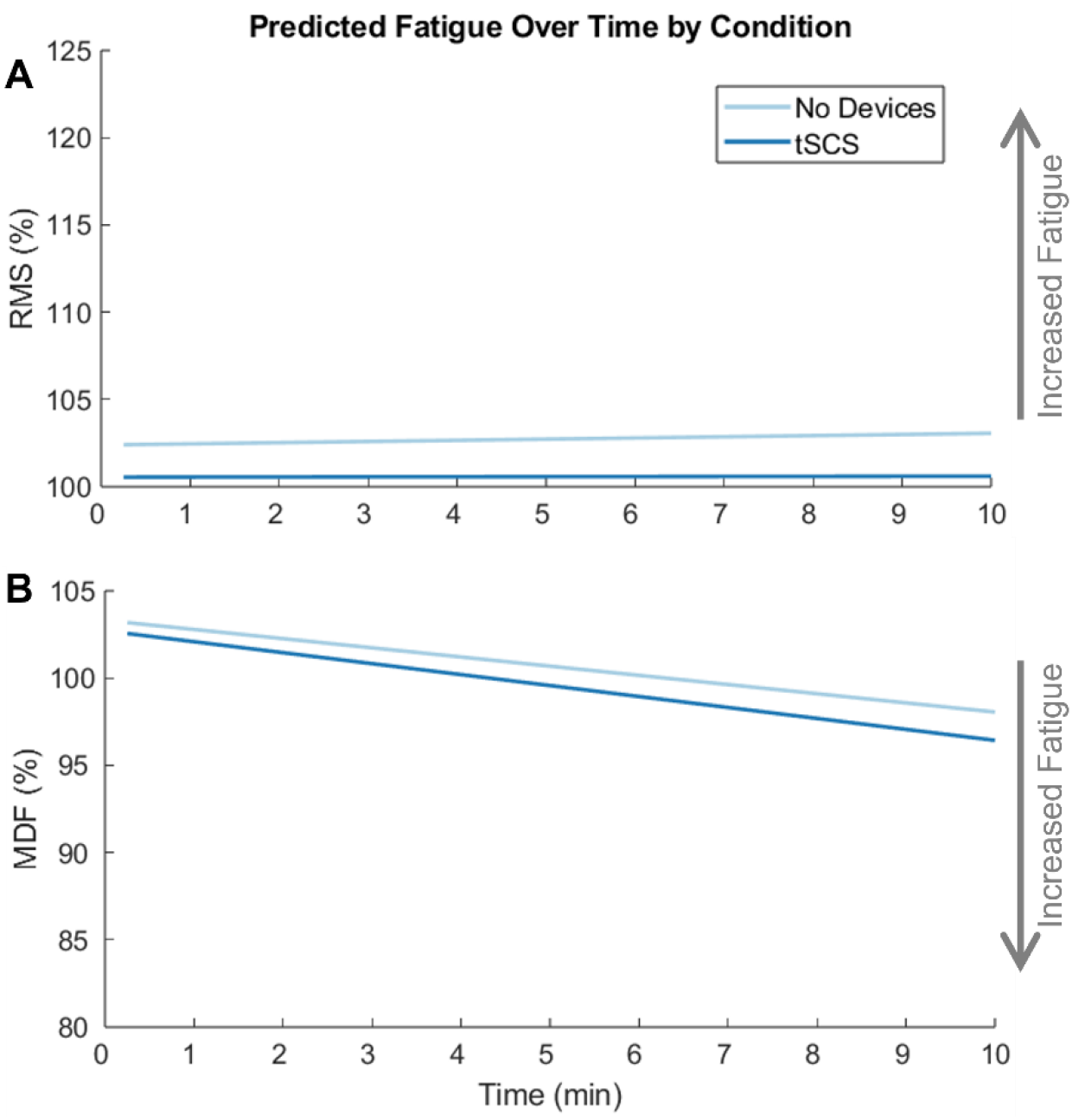
Myoelectric manifestations of muscle fatigue during ten minutes treadmill walking under two different conditions: No Device and tSCS only. A) The LMM for RMS of the EMG signal normalized to the peak RMS during the first 15 seconds of each condition. B) The LMM for MDF of the EMG signal, normalized to the MDF in the first 15 seconds of each condition. There were no significant differences between the tSCS and No Device condition for either RMS or MDF (p > 0.05).

Walking for 10 minutes with No Device led to no changes in RMS (p-value = 0.81) and a small but significant reduction in MDF (𝛽_3_ = -0.0088 %/s [-0.014, -0.0031]; p-value < 0.002). Walking with tSCS for ten minutes resulted in no significant differences in RMS or MDF compared to walking with No Device (RMS p-value = 0.88; MDF p-value = 0.68).

#### Fatigue during Pre-Post MVC

Despite the observed muscle fatigue within the first 5 minutes of the Exo condition, there were no significant changes in MVC force after twenty minutes of walking for any condition (Figure 5; p = 0.20). The force produced during the MVC task had a median increase after the Exo only condition of 6% (-15 to 34 Newtons, N), and decreased after training with No Device, tSCS only, and tSCS+Exo with reductions of 18% (-51 to 23 N), 3% (-15 to 41 N) and 9% (-43 to 7 N), respectively. Both RMS and MDF of the EMG signal increased slightly during the MVC after each walking trial, but the changes were similar between conditions (RMS p = 0.29; MDF p = 0.95). The EMG RMS during MVC increased by only 6%, 2%, 3%, and 7% after training with No Device, tSCS only, Exo only, and tSCS+Exo, respectively. The EMG MDF during MVC increased 6%, 6%, 7%, and 5% after trainingwith No Device, tSCS only, Exo only, and tSCS+Exo, respectively.

**Figure 5.**
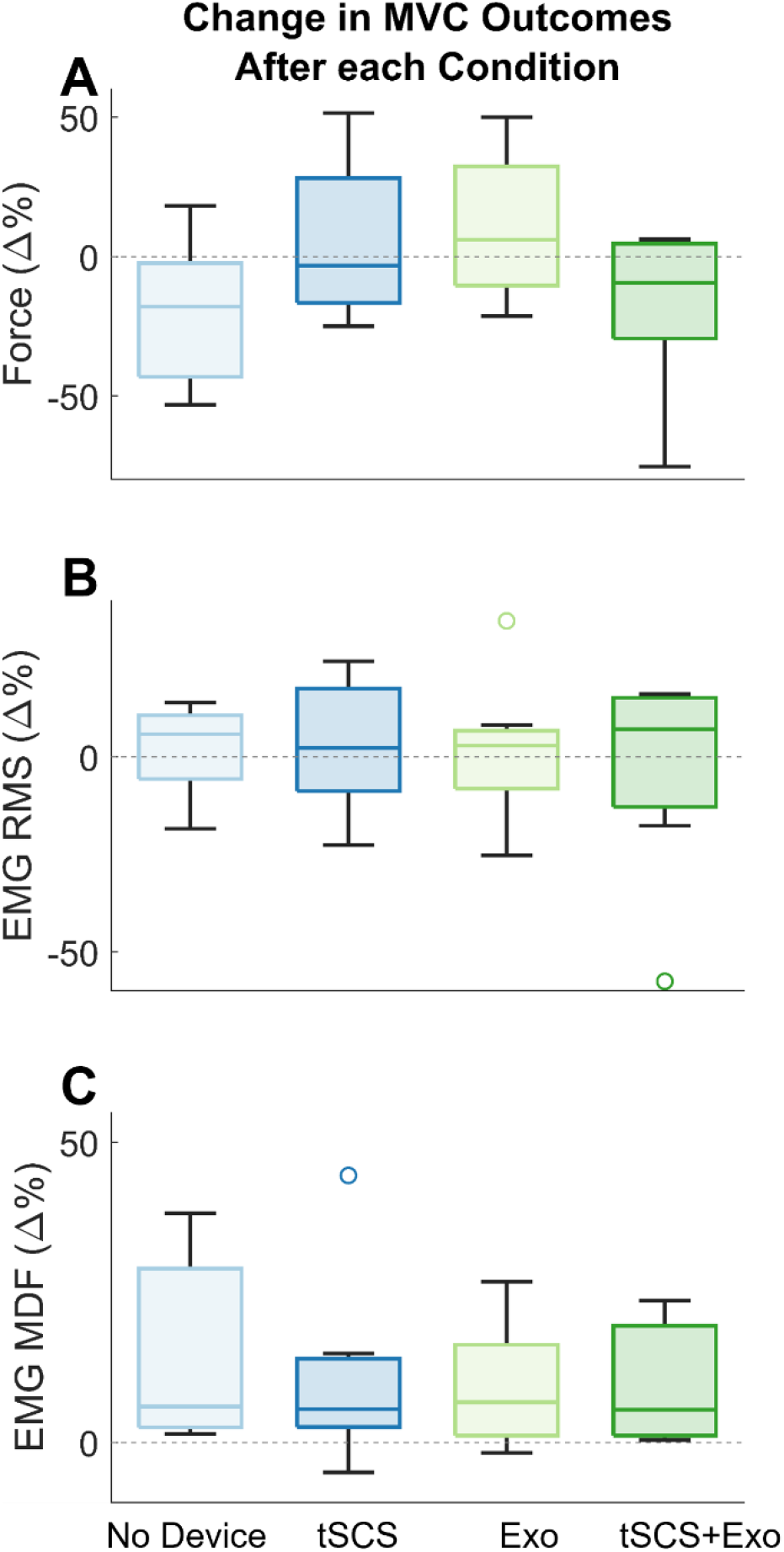
Percent change during plantarflexor MVC on the more-affected side in A) peak force, B) soleus EMG RMS and C) soleus EMG MDF after each walking condition.

## Discussion

As hypothesized, training with the Exo induced the greatest rate of muscle fatigue within the first five minutes of walking, as indicated with a simultaneous increase in soleus RMS and decrease in MDF. Soleus fatigue was not observed during the tSCS+Exo condition, suggesting that tSCS may reduce the impact of the Exo on muscle fatigue during training. Walking with No Device or with tSCS only were very similar in their rates of muscle fatigue, suggesting that five to ten minutes of treadmill training at a comfortable speed induced minimal myoelectric manifestations of muscle fatigue. Changes in the soleus RMS and MDF during MVC also did not indicate muscle fatigue.

Children with CP have greater muscle fatigue during five minutes of walking compared to typically developing (TD) peers. Eken et al., (2019) reported a significantly greater rate of fatigue via increased EMG RMS and decreased MDF in the gastrocnemius and soleus muscles on the more-affected side of children with CP compared to TD peers^29^. Their study evaluated overground walking, while here we utilized walking on a treadmill, which may offload the plantarflexors and put more demand on the quadriceps^30^. Parent et al., (2019) reported significant reductions in rectus femoris MDF across six minutes of treadmill walking in children with CP, but no change in other muscles^10^. Similarly, we did not observe simultaneous changes in RMS and MDF that would indicate muscle fatigue in the soleusduring five minutes of treadmill walking with No Device. The exoskeleton provides a dorsiflexion torque that counters the desired action at push-off, to encourage the user to increase plantarflexor engagement^18^. The increased rate of fatigue during training with the Exo suggests that the Exo successfully engaged the soleus and significantly increased the rate of fatigue. These findings are specifically for a resistive ankle exoskeleton. Assistive ankle exoskeletons have been shown to reduce muscle demand for children with CP and may result in reduced muscle fatigue^31^.

In ten consecutive minutes of walking with No Device and tSCS only, we observed a small but significant decline in MDF with no change in RMS. Changes in force output can also induce changes in RMS and MDF^27^. It is possible that in addition to some muscle fatigue, participants also had a simultaneous decline in force output that masked myoelectric manifestations of muscle fatigue in the RMS. We aimed to control for differences in force produced at the ankle by keeping walking speed consistent between walking conditions. But participants were allowed to use handrails for safety, which could have offloaded some bodyweight from the legs. Future work measuring force output either with an exoskeleton or instrumented treadmill could examine changes in force alongside fatigue.

We observed minimal changes in the EMG RMS and MDF during the MVC task performed before and after each walking condition. Prior work evaluating fifteen minutes of treadmill training has shown to reduce MVC force of the quadricep muscles by 11% in children with CP, with no change among TD peers^32^. In our study, we focused only on the soleus muscles, which did not indicate any fatigue with assessment of the MVC tasks alone. The increased sensory feedback when using either Exo or tSCS may improve coordination of muscles and increase the ability of the user to maximally engage in the MVC task, despite muscle fatigue^16,33^. Thus, future work should consider muscle fatigue and coordination of activity across multiple muscles to identify the possible mechanisms altering fatigue during walking, but not during the pre-post MVC tasks. Another possibility for no change during the MVC testing is that this task is not as physically demanding as walking for children with CP and occurs after the children have had a break from walking. Prior research has suggested that children with CP show similar and even reduced rates offatigue compared to nondisabled peers during tasks requiring maximal effort, but this maximal effort (i.e. force output) is also reduced^29,34^. One contributing factor to reduced maximal effort in children with CP is the potential activation of fewer muscle fibers during volitional maximal effort tasks compared to TD peers^35^. As a result, we may observe muscle fatigue more accurately during walking, where the demand on muscles could be higher than during the MVC task.

There are several limitations of this study. First, we evaluated the first five minutes of walking in our initial linear mixed effects model of all walking conditions, similar to prior studies on fatigue during walking in children with CP^10,28^. We were unable to include the conditions with the Exo for more than five minutes due to participants taking rest breaks and using the audiovisual feedback after the first five minutes to focus on one leg at a time^26^. Future research should consider alternative analysis methods to capture these additional cofounders and evaluate fatigue over longer time periods^36^. Second, we quantified fatigue in the soleus muscle, as the exoskeleton primarily targets the plantarflexors and covers the gastrocnemius. The soleus is generally considered the more fatigue resistant plantarflexor muscle^37^, so greater changes in fatigue may be observed in the gastrocnemius. The exoskeleton may also increase the engagement of other muscles, such as the rectus femoris. Additional EMG processing will be needed to remove signal artifact from tSCS for more proximal muscles^38^. Finally, MVC was quantified with a manual dynamometer with resistance to plantarflexion provided by a researcher. Measuring MVC with a manual dynamometer is a common practice^18,29,39,40^, but use of an isokinetic dynamometer, such as a Biodex system, would produce more accurate force measurements. Further, a direct recording of forces produced during walking could clarify changes in RMS that may also be due to force output.

In conclusion, walking with an Exo increased the rate of muscle fatigue during the first five minutes of treadmill walking compared to walking with No Device. The addition of tSCS reduced fatigue in the soleus muscle and likely increased force output and, thus, engagement with the device. These results suggest that tSCS may be a viable tool for reducing muscle fatigue during training to maximize rehabilitation benefits of physical therapy for children with CP.

## Data Availability

All data produced in the present study are available upon reasonable request to the authors.

## Data Availability

The datasets generated during and/or analyzed during the current study are available from the corresponding author on reasonable request.

## Acknowledgements

The authors thank the children and their families for the time they dedicated to the research. We also thank the fellow researchers who supported our data collection visits and preliminary data analysis, especially Katherine Green and Yusuke Maruo. Additional researchers and clinicians who supported data collection included Roozbeh A. Katiraie, Michael Hall, Kathleen Landwehr-Prakel, and Lindsey Jouett.

## Author Contributions

All authors conceived of and designed the research. C.R.D., V.M.L., and S.R.S. performed data collection. C.R.D., V.M.L., and K.M.S. carried out all data analysis and interpreted experimental results. C.R.D. and V.M.L. prepared figures and drafted the manuscript. All authors edited and revised the manuscript and approved the final version.

### Funding

This work was supported by Seattle Children’s Hospital CP Research Pilot Study Fund 2022 Award, NSF Graduate Research Fellowship Program Award DGE-1762114, and the National Institute of Neurological Disorders and Stroke of the National Institutes of Health under award number R01NS091056.

## References

1. Bolster, E. A. M., Balemans, A. C. J., Brehm, M. A., Buizer, A. & Dallmeijer, A. J. Energy cost during walking in association with age and body height in children and young adults with cerebral palsy. Gait Posture 54, 119–126 (2017).

2. Waters, R. L. & Mulroy, S. The energy expenditure of normal and pathologic gait. Gait Posture 9, 207–231 (1999).

3. Rose, J., Gamble, J. G., Burgos, A., Medeiros, J. & Haskell, W. L. Energy expenditure index of walking for normal children and for children with cerebral palsy. Dev Med Child Neurol 32, 333–340 (1990).

4. Jahnsen, R., Stanghelle, J. K. & Holm, I. Fatigue in adults with cerebral palsy in Norway compared with the general population. Dev Med Child Neurol 45, 296–303 (2003).

5. Hilberink, S. R. et al. Health issues in young adults with cerebral palsy: Towards a life-span perspective. J Rehabil Med 39, 605–611 (2007).

6. Dominici, N. et al. Systematic Review of Fatigue in Individuals With Cerebral Palsy. Front Hum Neurosci 15, 1–16 (2021).

7. Gandevia, S. C. Spinal and Supraspinal Factors in Human Muscle Fatigue. Physiol Rev 81, 1725–1789 (2001).

8. Bigland Ritchie, B., Johansson, R., Lippold, O. C. J. & Woods, J. J. Contractile speed and EMG changes during fatigue of sustained maximal voluntary contractions. J Neurophysiol 50, 313–324 (1983).

9. De Luca, C. J. Myoelectrical manifestations of localized muscular fatigue in humans. Crit Rev Biomed Eng 11, 151–279 (1984).

10. Parent, A. et al. Muscle fatigue during a short walking exercise in children with cerebral palsy who walk in a crouch gait. Gait Posture 72, 22–27 (2019).

11. Eken, M. M. et al. Squat test performance and execution in children with and without cerebral palsy. Clinical Biomechanics 41, 98–105 (2017).

12. Moreau, N. G., Li, L., Geaghan, J. P. & Damiano, D. L. Fatigue Resistance During a Voluntary Performance Task Is Associated With Lower Levels of Mobility in Cerebral Palsy. Arch Phys Med Rehabil 89, 2011–2016 (2008).

13. Stackhouse, S. K., Binder-Macleod, S. A. & Lee, S. C. K. Voluntary Muscle Activation, Contractile Properties, and Fatigability in Children With and Without Cerebral Palsy. Muscle Nerve 31, 594–601 (2005).

14. Xu, Y. et al. Rehabilitation Effects of Fatigue-Controlled Treadmill Training After Stroke: A Rat Model Study. Front Bioeng Biotechnol 8, 1–17 (2020).

15. Ma, J., Chen, H., Liu, X., Zhang, L. & Qiao, D. Exercise-Induced Fatigue Impairs Bidirectional Corticostriatal Synaptic Plasticity. Front Cell Neurosci 12, 1–14 (2018).

16. Conner, B. C., Schwartz, M. H. & Lerner, Z. F. Pilot evaluation of changes in motor control after wearable robotic resistance training in children with cerebral palsy. J Biomech 126, 110601 (2021).

17. Conner, B. C., Luque, J. & Lerner, Z. F. Adaptive ankle resistance from a wearable robotic device to improve muscle recruitment in cerebral palsy. Ann Biomed Eng 48, 1309–1321 (2020).

18. Conner, B. C., Remec, N. M., Orum, E. K., Frank, E. M. & Lerner, Z. F. Wearable adaptive resistance training improves ankle strength, walking efficiency and mobility in cerebral palsy: A pilot clinical trial. IEEE Open J Eng Med Biol 1, 282–289 (2020).

19. DeVol, C. R. et al. Effects of spinal stimulation and short-burst treadmill training on gait biomechanics in children with cerebral palsy. Gait Posture 118, 25–32 (2025).

20. Hastings, S. et al. A pilot study combining noninvasive spinal neuromodulation and activity-based neurorehabilitation therapy in children with cerebral palsy. Nat Commun 13, 1–7 (2022).

21. Gad, P. et al. Transcutaneous spinal neuromodulation reorganizes neural networks in patients with cerebral palsy. Neurotherapeutics 18, 1953–1962 (2021).

22. Solopova, I. A. et al. Effects of spinal cord stimulation on motor functions in children with cerebral palsy. Neurosci Lett 639, 192–198 (2017).

23. Shrivastav, S. R. et al. Transcutaneous spinal stimulation and short-burst interval treadmill training in children with cerebral palsy: a pilot study. IEEE Trans Biomed Eng 72, 1–12 (2025).

24. Sachdeva, R., Girshin, K., Shirkhani, Y., Gad, P. & Edgerton, V. R. Combining spinal neuromodulation and activity based neurorehabilitation therapy improves sensorimotor function in cerebral palsy. Frontiers in Rehabilitation Sciences 4, 1–5 (2023).

25. Keller, A. et al. Noninvasive spinal stimulation safely enables upright posture in children with spinal cord injury. Nat Commun 12, 1–11 (2021).

26. Spomer, A. M., Conner, B. C., Schwartz, M. H., Lerner, Z. F. & Steele, K. M. Audiovisual biofeedback amplifies plantarflexor adaptation during walking among children with cerebral palsy. J Neuroeng Rehabil 20, 1–17 (2023).

27. Cifrek, M., Medved, V., Tonković, S. & Ostojić, S. Surface EMG based muscle fatigueevaluation in biomechanics. Clinical Biomechanics 24, 327–340 (2009).

28. Eken, M. M. et al. Lower limb muscle fatigue during walking in children with cerebral palsy. Dev Med Child Neurol 61, 212–218 (2019).

29. Eken, M. M., Dallmeijer, A. J., Houdijk, H. & Doorenbosch, C. A. M. Muscle fatigue during repetitive voluntary contractions: A comparison between children with cerebral palsy, typically developing children and young healthy adults. Gait Posture 38, 962–967 (2013).

30. Lee, S. J. & Hidler, J. Biomechanics of overground vs. treadmill walking in healthy individuals. J Appl Physiol 104, 747–755 (2008).

31. Lerner, Z. F., Damiano, D. L. & Bulea, T. C. The effects of exoskeleton assisted knee extension on lower-extremity gait kinematics, kinetics, and muscle activity in children with cerebral palsy. Sci Rep 7, 1–12 (2017).

32. Vitiello, D. et al. Walking-induced muscle fatigue impairs postural control in adolescents with unilateral spastic cerebral palsy. Res Dev Disabil 53–54, 11–18 (2016).

33. Edgerton, V. R., Hastings, S. & Gad, P. N. Engaging Spinal Networks to Mitigate Supraspinal Dysfunction After CP. Front Neurosci 15, 1–5 (2021).

34. Leunkeu, A. N., Keefer, D. J., Imed, M. & Ahmaidi, S. Electromyographic (EMG) Analysis of Quadriceps Muscle Fatigue in Children With Cerebral Palsy During a Sustained Isometric Contraction. J Child Neurol 25, 287–293 (2010).

35. Rose, J. & McGill, K. C. Neuromuscular activation and motor-unit firing characteristics in cerebral palsy. Dev Med Child Neurol 47, 329–336 (2005).

36. DeVol, C. R. et al. Effects of interval treadmill training on spatiotemporal parameters in children with cerebral palsy: A machine learning approach. J Biomech 177, 1–8 (2024).

37. Cronin, N. J., Avela, J., Finni, T. & Peltonen, J. Differences in contractile behaviour between the soleus and medial gastrocnemius muscles during human walking. Journal of Experimental Biology 216, 909–914 (2013).

38. Kim, M. et al. A novel technique to reject artifact components for surface EMG signals recorded during walking with transcutaneous spinal cord stimulation: A pilot study. Front Hum Neurosci 15, 1–14 (2021).

39. O’Brien, S. M., Carroll, T. J., Barber, L. A. & Lichtwark, G. A. Plantar flexor voluntary activation capacity, strength and function in cerebral palsy. Eur J Appl Physiol 121, 1733–1741 (2021).

40. Tedroff, K., Knutson, L. M. & Soderberg, G. L. Co-activity during maximum voluntary contraction: a study of four lower-extremity muscles in children with and without cerebral palsy. Dev Med Child Neurol 50, 377–381 (2008).

